# A randomized controlled trial of patient navigators for post-hospital discharge diabetic foot ulcer care: the Comprehensive Assistance and Resources for Effective Diabetic Foot Navigation (CARE-D-Foot-Nav) study protocol

**DOI:** 10.1101/2025.11.25.25340976

**Authors:** Marcos C. Schechter, Elizabeth C. Rhodes, Hui Shao, Siham Ahmed, Kytabia Colquitt, Nataliya Y. Chaudhry, Anne L. Dunlop, Jesica Flores, Manuel Garcia-Toca, Vidhi Javia, Ameeta S. Kalokhe, Aadhiti Manoj, Candace Meadows, Natalie Meriwether, Jessica M. Sales, Nazanin Soleimanmanesh, Gabriel Santamarina, Neena K. Smith-Bankhead, Guillermo Umpierrez, Christopher R. Ramos, Emily Watson, Limin Peng, Maya Fayfman

## Abstract

**Background:** Diabetic foot ulcers (DFUs) are a leading cause of limb loss globally, and DFU-related amputation rates are increasing in the United States. Multidisciplinary care improves DFU healing and prevents amputations. This strategy encompasses 4 key pillars: optimizing glycemic control, managing wounds, treating vascular disease, and addressing infections. However, the implementation of these pillars is fraught with fragmented and delayed care impeding effective wound healing and leading to amputations. Patient navigation improves chronic disease outcomes, including diabetes, but have not been tested for DFU care.

**Methods/Design:** The Comprehensive Assistance and Resources for Effective Diabetic Foot Navigation (CARE-D-Foot-Nav) is a multicomponent patient navigator program tailored to address individual and structural obstacles to would healing. Adults hospitalized with a DFU (n=270) will be randomized 1:1 to receive the CARE-D-Foot-Nav intervention or usual care. The primary outcome is complete DFU healing at 20-weeks post-hospital discharge adjudicated by independent reviewers blinded to treatment assignment. Secondary outcomes include patient-reported measures and economic outcomes.

**Discussion:** CARE-D-Foot-Nav is the first randomized controlled trial to evaluate patient navigation for DFU care. If the patient navigator program improves outcomes, it could transform care delivery for people with DFUs and reduce preventable limb loss.

**Trial registration:** ClinicalTrials.gov NCT07223268

## INTRODUCTION

Up to one-third of the 38 million people living with diabetes in the United States (US) will develop a diabetic foot ulcer (DFU) in their lifetime^1,2^. DFUs are extremely costly amounting to $78 billion per year in the US and lead to a poor quality of life^1^. Poor DFU healing is the leading cause of non-traumatic preventable amputations in the US^3^ and globally^4^. Unfortunately, despite numerous advances in diabetes care therapeutics^5,6^, the number of diabetes-related lower-extremity amputations is increasing in the US and over 150,000 amputations occurred in this country in 2020^3^. Multidisciplinary care improves DFU healing and prevents amputations^7^. This strategy encompasses 4 key pillars: optimizing glycemic control, managing wounds, treating vascular disease, and addressing infections^8–10^. However, the real-world implementation of these pillars is often fragmented and delayed, impeding wound healing leading to amputations ^11–15^.

Here, we report the protocol of a randomized controlled trial designed to investigate if a multicomponent patient navigator program (CARE-D-Foot-Nav) tailored to high-risk populations that addresses individual and structural obstacles to care improves DFU healing^15^. Patient navigators are healthcare personnel who mitigate obstacles to care and improve chronic diseases outcomes. A systematic review of randomized controlled trials (RCTs) found that navigators improved outcomes in 7 of the 8 diabetes-specific studies, including in high-risk populations ^16–23^. DFUs, however, are a unique diabetes complication, and effective treatment requires coordination across numerous specialties, making it susceptible to care fragmentation^8–10^. Existing multidisciplinary DFU programs are not equipped with tools to mitigate the impact of individual drivers of adverse health outcomes such as transportation limitations and poor health literacy^7^ Notably, no RCTs to date have tested a patient navigator specifically for DFU care^24^.

The CARE-D-Foot-Nav DFU navigator program was informed by extensive preliminary data showing suboptimal implementation of the 4 pillars of care at the healthcare system where the trial will be conducted ^11,14,25,26^, as well as community input and a pilot navigator trial^27^. The CARE-D-Foot-Nav DFU navigator’s main goal is to assist participants in attaining the 4 pillars of care. To achieve this, the navigator will maintain close contact with patients through weekly encounters (phone or in person) and support them through their healing journey with resources tailored to their unique needs (e.g., support diabetes and wound management, transportation reimbursement, and coordinate outpatient appointments). To capture those at highest risk of complications, we will enroll adults hospitalized with a DFU and randomize them to CARE-D-Foot-Nav *vs* usual care for 20 weeks post-discharge. We hypothesize that CARE-D-Foot-Nav will improve linkage to multidisciplinary care with a focus on the pillars of DFU care and improve DFU healing. This RCT will enroll 270 participants in a high DFU volume hospital in Atlanta, Georgia^11,26^ over 4 years. Implementation will be guided by the RE-AIM framework^28^, and the Consolidated Framework for Implementation Research (CFIR) will be used to identify barriers obstacles and facilitators to implementation outcomes such as fidelity and acceptability^29,30^.

## OBJECTIVES

This pragmatic, open-label randomized controlled trial will investigate the effectiveness of a patient navigator program (CARE-D-Foot-Nav) for adults hospitalized with DFUs to achieve a primary outcome of DFU healing at 20 weeks post-hospital discharge, defined as complete re-epithelialization adjudicated by blinded, independent reviewers. This single-center trial will be conducted at Grady Memorial Hospital in Atlanta, Georgia, one of the largest public safety net hospitals in the US with over 250 people with DFU admitted annually^11,26^. In addition to the primary outcome of 20-week healing, we will investigate a range of secondary clinical and patient-reported outcomes. We will also evaluate the implementation fidelity and acceptability of the navigator program using mixed-methods and perform a cost-effectiveness analysis of the CARE-D-Foot-Nav program. The RCT protocol and results report will follow the SPIRIT^31^ and CONSORT^32^ statements as appropriate.

## METHODS

### Study design

This single-center RCT will randomize 270 adults hospitalized with a DFU at Grady Memorial Hospital, Atlanta, GA, 1:1 to CARE-D-Foot-Nav *vs* usual care for 20 weeks post-hospital discharge (Figure 1). Due to the high risk of ulcer recurrence^33^, we will conduct a follow-up visit at week 22 to confirm wound healing. Our overarching hypotheses are that (1) patient navigators increase access to the 4 pillars of multidisciplinary DFU care and thus (2) improve DFU healing (primary outcome). To test residual effects after the intervention period, we included an observational period from weeks 20 to 52 post-hospital discharge in which both arms receive usual care, and we will evaluate differences in rates of amputations and healthcare utilization. Potentially eligible inpatients will be screened and enrolled during a run-in phase, the period between enrollment and hospital discharge. Participants who become ineligible during the run-in phase will be excluded and classified as screen failures. Participants who complete the run-in phase will be randomized within two business days post-hospital discharge using block randomization prepared by the trial statistician (LP) to achieve balanced allocation across the two study arms.

**Figure 1:**
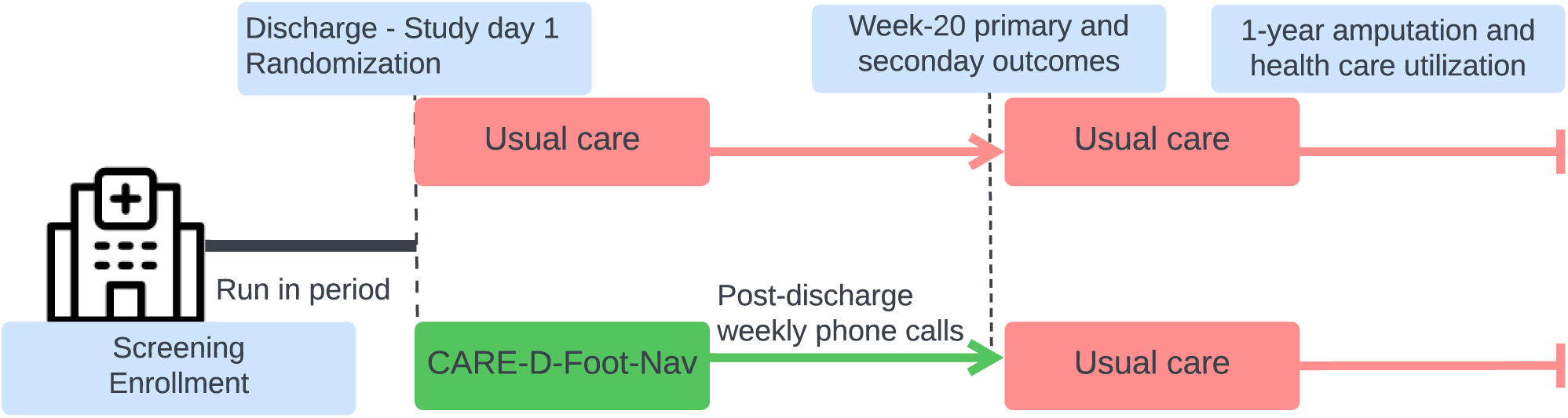
study design.

### Participants eligibility criteria

Consistent with the pragmatic nature of the trial, we will broadly include adults (≥18 years old) with diabetes hospitalized for any reason who have a full-thickness DFU, defined as a wound below the ankle through the dermis, or a single toe amputation. We will exclude people who are unable to understand the nature and scope of the study, enrolled in another clinical trial, planned for discharge to an acute or long-term care facility, or planning to receive all outpatient DFU care outside of Grady Memorial Hospital. Amputations of any level prior to hospital admission are not an exclusion criterion and participants who undergo a single toe amputation (including those with primary closure) during the hospitalization are eligible for inclusion. People who undergo an amputation of ≥2 toes during the index hospitalization will be considered screen failures. We will also exclude those with a Society for Vascular Surgery Wound, Ischemia, foot Infection (WIfI) score of 4, as healing and limb salvage are unlikely^34–36^.

## INTERVENTIONS

### Patient navigator group (intervention)

The overarching goal of the navigator is to help patients access the 4 key pillars of DFU care to optimize the chances of DFU healing (table 1). Participants randomized to CARE-D-Foot-Nav will be assigned a dedicated DFU patient navigator. Based on community input, the navigators will conduct 30-60-minute encounters either by phone or in person at least once a week during the 20-week program. Additional contact may be warranted to coordinate appointments and transportation. Participants can call the navigator with DFU-related concerns anytime during business hours. Based on community feedback, the navigators will receive Trauma Informed Care (TIC) and motivational interviewing training. The TIC training purpose is to introduce navigators to a healing-centered care approach to improve service delivery and patient care outcomes. The terminology used in the sessions was purposefully transitioned from “trauma-informed care” to “healing-centered care” throughout the training to be responsive to changes in the field that called for a shift from a focus on trauma and treating traumatic events to a focus on healing. The overarching goal of the training is to empower individuals to participate and lead changes to improve their own well-being, centering on both individual and collective healing. Motivational interviewing is a collaborative, compassionate communication style that helps patients find their own internal motivation for behavior change, leading to better health outcomes^37^.

**Table 1:** the CARE-D-Foot-Nav patient navigator program.

| Domain | Components |
| --- | --- |
| **1-Pillars of diabetic foot ulcer care (frequency: weekly)** |  |
| Glycemic control | (1a) Review treatment plan as outlined by treating clinicians<br>(1b) **Escalation of care trigger:** Symptomatic hyper- or hypoglycemia |
| Wound management | (2) Review wound care plan (3) education on rationale for DFU treatments and natural history (4) activity recommendations, (5) inquire about wound worsening, (6) assess need for and provide wound care supplies<br>(7) **Escalation of care trigger:** Patient reported new or worsening signs of infection (self-reported or observed redness, drainage, increase in size) |
| Infection management | (8a) Reinforce antibiotic therapy plan, if applicable. (8b) **Escalation of care trigger:** Antibiotic intolerance or issues infusing antibiotics if IV antibiotics were prescribed |
| PAD management | (9) Review peripheral artery disease (PAD) diagnostic test orders and schedule tests, if needed |
| **2- 30-day post discharge access to pillars of care (weekly for first 4 weeks post discharge)** |  |
| Pillars of care | (10) Attainment of the 4 pillars of care with clinical visits (as indicated) for a) diabetes management b) wound care c) infectious diseases. d) vascular within 30 days of hospital discharge<br>This applies to infectious diseases and vascular surgery only if the respective services were recommended post-hospital discharge follow-up. |
| **3-Tools to improve access to diabetic foot ulcer care and emotional support (frequency: weekly)** |  |
| Outpatient care coordination | (11) Remind participant of appointments, (10) offer to assist with consolidating schedule, (11) offer to attend DFU outpatient clinic visits with the patient |
| Transportation assistance | (12) Offer Rideshare, parking vouchers, and/or fuel funds |
| Language-concordant care | (13) Language concordant navigator for Spanish-speaking patients |
| Peer support | (14) Assess interest in diabetic foot ulcer peer support group meetings and facilitate peer support group meetings for interested participants |
| Support if amputations are recommended | (15) Offer to meet with the patient and help understand options and care if the medical team recommends an amputation, if applicable |
| Spiritual Support | (16) Assess interest in spiritual support and (16a) hospital's chaplaincy service referral, if applicable |
| **4-Depression screening (frequency: at program enrollment and midway between weeks 11-13)** |  |
| Depression screening (PHQ-2 and PHQ-9) | (17) Screen for depression (17a) counselling and/or diabetes support group referral, if appropriate<br>(17b) **Escalation of care trigger:** Participant reports suicidal or homicidal ideation |
| **5-Access to existing services and addressing individual drivers of adverse health outcomes (frequency: at program enrollment and midway between weeks 11-13)** |  |
| Tobacco use screening | (18) Assess for tobacco use and (18a) make tobacco cessation program referral and number for smoking cessation programs, if applicable |
| Food insecurity screening | (19) Assess for food insecurity and (19a) make food bank referral, if applicable |
| Alcohol use disorder | (20) Alcohol use disorder screening and (20a) Alcohol cessation program referral, if applicable |
| Financial resource strain | (21) Assess for financial resource strain and (21a) social work referral, if applicable |
| Housing instability | (22) Assess for housing instability and (22a) housing navigator referral, if applicable |

### First CARE-D-Foot-Nav navigator encounter (post-randomization)

The first patient navigator encounter will occur after randomization during hospitalization up to 2 business days of discharge. The navigator will conduct an intake interview to personalize the program to the participant’s needs, including communication preferences (phone, in person, or both), obtain alternative contacts (in case of missed encounters), and determine which program components they are interested in. This extended session will also include a review of discharge medications and recommendations. The navigator will ensure the patient is aware of all recommended appointments and assist with scheduling if needed.

#### Four pillars of diabetic foot ulcer (DFU) care components

##### 1. Glycemic control

<u>Justification</u>: Our group and others have shown that optimized glycemic control promotes wound healing, including for DFU^38–40^. <u>Intervention</u>: At each encounter, the navigator will assess participants’ glycemic control by self-monitoring of blood glucose and reinforce diabetes education and self-efficacy through connecting participants to available resources at Grady. <u>Escalation of care</u>: If a participant reports severe hypoglycemia (defined as an episode of low blood sugar with altered mental and/or physical status that requires assistance from another person for recovery) or hyperglycemia (prolonged hyperglycemia with BG of >350 over 24 hours or symptoms of severe hyperglycemia (nausea, vomiting, abdominal pain, and dehydration) despite at home treatment with standard of care, the navigator will contact the treating provider (clinical nurse, endocrinologist, or primary care doctor) or assist the patient in calling the nurse advice line to coordinate next steps. Patients with diagnosed type 1 diabetes will be advised to call the navigator if they are out of insulin and need urgent refills.

##### 2. Wound management

<u>Justification</u>: Wound debridement and offloading improve DFU healing^1,41–43^. Participants in the focus group reported not understanding the rationale for wound debridement, which led to medical mistrust. <u>Intervention</u>: The navigator will streamline and promote participants’ access to care and will also reinforce provider recommendations for wound care, offloading, footwear, and activity limitations, help participants understand the rationale for DFU treatments including debridement, and assess the need for and provide basic wound care supplies if not covered by their insurance. <u>Escalation of care:</u> When participants report worsening signs/symptoms of DFU, the navigator will ask them to send a picture of their foot and will contact the wound care provider (surgical or wound care) to review the images and determine an action plan.

##### 3. Infection treatment

<u>Justification</u>: DFU infection is usually the terminal event before limb loss and requires complex treatment plans^10,44^. <u>Intervention</u>: For participants with DFU infections, the navigator will review antibiotic management to ensure they are taking medications as directed by their infectious diseases provider. <u>Escalation of care</u>: For participants reporting antibiotic adverse effects, the navigator will contact the infectious diseases provider to determine if any interventions are needed.

##### 4. PAD management

<u>Justification</u>: PAD assessment and treatment are key for DFU healing^45^. Thus, guidelines recommend that all people with DFUs should have a battery of non-invasive tests for PAD that include toe brachial index, toe pressure, and pulse waveforms, collectively referred to as pulse volume recordings^9^. <u>Intervention</u>: The navigator will ensure all participants undergo pulse volume recordings if not completed within the past six months, following a vascular procedure, or if otherwise recommended by a provider. The navigator will assist with the coordination of invasive testing, such as angiography (when recommended by vascular surgeons). The navigator will contact providers to order testing if a referral is not already in place.

#### Escalation of Care protocol for glycemic control, wound care, and infection treatment

The navigator will contact the relevant provider or call the hospitals’ nurse advice line to advise on one or more of the following next steps: 1) adjust therapy; 2) schedule earlier follow-up visits; 3) recommend Emergency Room evaluation; or 4) no intervention. The navigator will then assist with implementation. If the provider cannot be reached, the principal investigators will assist with escalation of care.

#### Components to improve access to the four pillars of DFU care and emotional support

##### 1. Outpatient care coordination

<u>Justification</u>: We and others have found that patients report difficulty attaining appointments with the multiple providers required for DFU care. Focus group participants corroborated these concerns and expressed interest in a patient navigator to assist with continuity of care. <u>Intervention</u>: For clinical visits relating to the 4 pillars of DFU care, the navigator will facilitate scheduling (e.g., consolidate visits to a single day when possible and reschedule to fit participants’ needs) and call participants with appointment reminders one day prior. Grady policy states that all patients hospitalized with a DFU should have at least one diabetes-related (primary provider or endocrinologist) and one wound-related (surgical or wound care service) visit within 2 weeks of hospital discharge. No referral is needed for these services. The navigator will work with participants to ensure they are scheduled within 2 weeks of discharge. Navigators will offer to attend clinic visits with participants to assist with continuity of care, social support, and education.

##### 2. Transportation assistance

<u>Justification</u>: Transportation is a common barrier to chronic disease care in the US^46^, including for people with DFUs in Georgia^47,48^. <u>Intervention</u>: Participants will receive up to $500 over 20 weeks for transportation to DFU-related appointments that may be used for ridesharing, parking, or fuel depending on their needs; $500 is estimated to cover ∼10 round trip rideshares.

##### 3. Language-concordant care

<u>Justification</u>: Language concordant care improves outcomes such as glycemic control^49^. Spanish speaking patients account for 10% of Grady’s population, the second most common language following English. <u>Intervention</u>: Participants can select a navigator who is a fluent Spanish speaker.

##### 4. Peer support

Justification: Focus group participants expressed interest in DFU peer support meetings Intervention: The navigator will invite patients to attend monthly peer support meetings where they will be able to share personal experiences with DFU care.

##### 5. Support at the point of critical DFU care decision points

<u>Justification</u>: Amputations are the most feared DFU complication^50,51^, and our focus group participants expressed that this was a time where support from someone who knows them was most needed. <u>Intervention</u>: Navigators will provide support when amputations are recommended, including during hospital re-admissions. Participants who undergo amputation will remain in the navigator program to assess secondary outcomes (e.g., social support).

##### 6. Depression screening

<u>Justification</u>: Depression is common among people with DFUs and can adversely impact outcomes^52,53^. <u>Intervention</u>: Navigators will screen for depression with the PHQ-2 and, if positive, administer the PHQ-9^54^. If the PHQ-9 score is ≥10, the navigator will help the participant access Grady and community mental health resources. <u>Escalation of care</u>: If a participant reports suicidal or homicidal ideation, the navigator will contact the MPIs who will deploy emergency services if needed.

##### 7. Improve utilization of existing resources

<u>Justification</u>: Many health systems, including Grady, have resources in place that can benefit people with DFUs (e.g., smoking cessation programs, food banks, chaplaincy service), but patients and providers may not be aware of how to access them. <u>Intervention</u>: Navigators will screen participants for eligibility and interest in available resources and provide guidance on how to access them.

##### 8. Participants who do not have a mobile phone

Based on previous surveys, approximately 95% of patients receiving care in the Diabetes Center at Grady own a smartphone. If a participant randomized to CARE-D-Foot-Nav does not own a camera equipped mobile phone, we will provide phones that will be returned upon completion of the intervention.

##### 9. Rescue for participants who miss CARE-D-Foot-Nav encounters for ≥2 consecutive weeks

The navigator will reach out to the alternate contact if one was provided. Participants who miss ≥4 consecutive weeks will be withdrawn from the program.

The CARE-D-Foot intervention processes, explanatory mechanisms, and their measurements depicted in the research implementation logic model (figure 2)^55^

**Figure 2:**
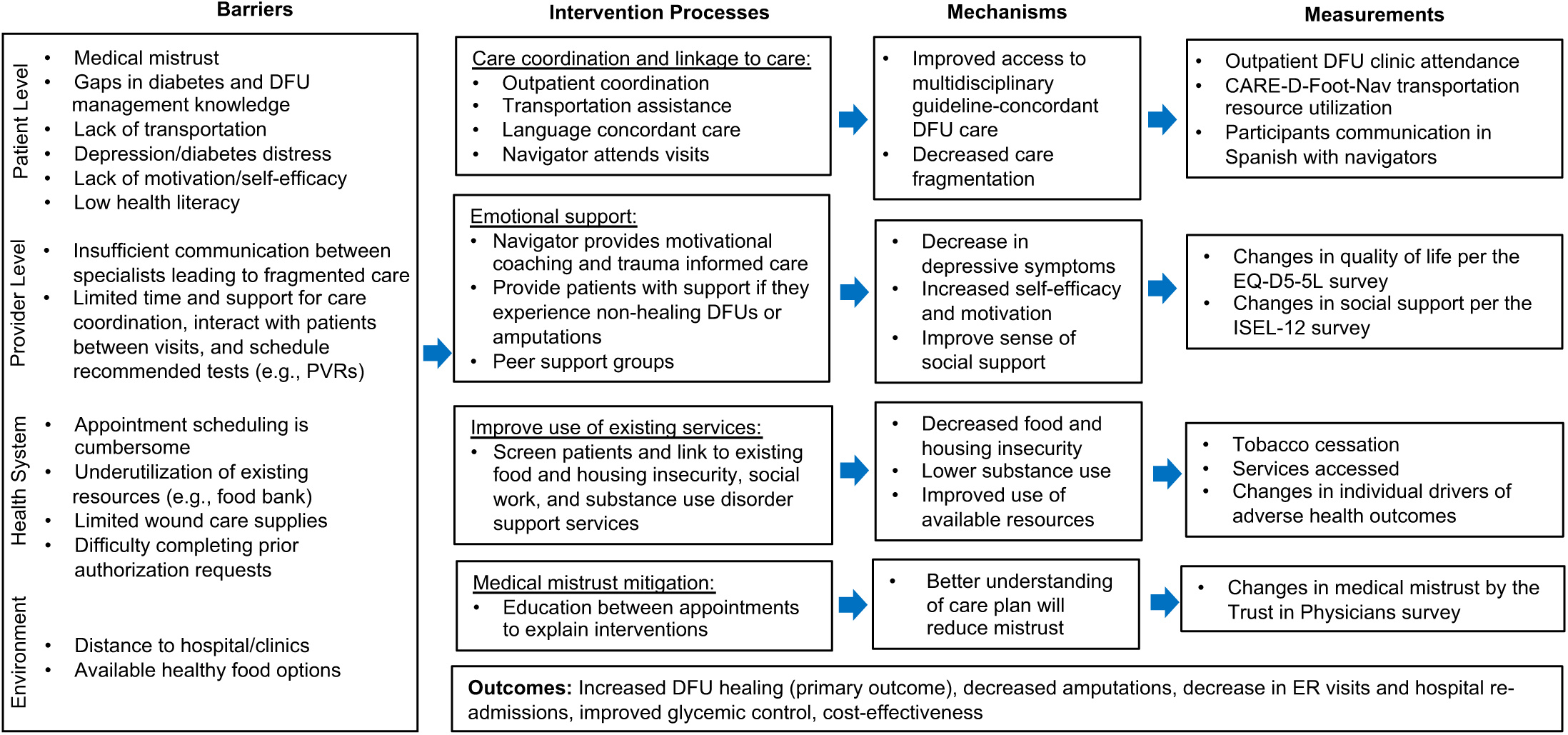
The CARE-D-Foot-Nav conceptual framework.

#### Control

Participants randomized to usual care will complete the social determinants of health surveys and receive a DFU Resource Guide. Research coordinators will conduct follow-up calls at 4-, 8-, 12-, 16-, and 20-week post-hospital discharge to support retention in the trial, and, for ethical reasons, the coordinator will remind participants of upcoming clinic appointments and/or attempt to link them back to DFU care if no appointments are scheduled. To measure time to healing, all participants (navigator and control arm) will be asked to send photos of their ulcer and answer a short survey of self-reported wound status at each 4-week interval.

### DATA COLLECTION AND MANAGEMENT

Data will be collected using a standardized REDCap case report forms^56^. We will record all patient navigator and control encounters to assess fidelity to the navigator intervention and control protocols. Additionally, we will obtain baseline DFU images at study enrollment and at 4-week intervals up to week 20 (+/- 7 days) post-hospital discharge for the primary outcome of DFU healing measurement, and 2 weeks (11 to 21 days) after the week 20 image was obtained to confirm the wound healing status, which is a secondary outcome. We will use an Emory University-based HIPAA-compliant server to store encounter audio and image files.

### OUTCOMES

We will use elements of the RE-AIM (Reach, Effectiveness, Adoption, Implementation, and Maintenance) framework to guide our evaluation of CARE-D-Foot-Nav as described in table 2.

**Table 2:** evaluation of the CARE-D-Foot-Nav program by RE-AIM dimension.

|  |
| --- |
| Reach |
| Report CONSORT diagram<br>Compare characteristics of the trial participants to those of the overall population with DFUs |
| Effectiveness |
| Key clinical outcomes (1) healing rate and (2) attainment of the four pillars of DFU care<br>Broader outcomes measurement, including patient reported outcomes |
| • Measure robustness across subgroups<br>• Measure attrition: report CARE-D-Foot-Nav program week-20 dropout rates |
| Implementation |
| • Fidelity: participant adherence to weekly navigator encounters<br>• Fidelity: consistency of implementation across navigators and stratified by participant characteristics<br>• Qualitative participant, navigator, and provider interviews to evaluate limitations and facilitators to fidelity and acceptability<br>• Cost-effectiveness analysis<br>• Program adaptations, including from community advisor report |

#### Primary outcome

the RCT primary outcome is healing 20-week post-hospital discharge (day 140 +/-7 days post-hospital discharge). DFU healing is defined as complete re-epithelialization of the wound as adjudicated by independent, blinded reviewers, in line with DFU RCT best practices^57^. We will take photographs of the wounds at enrollment, every 4 weeks up to week 20, and at the week 22 confirmatory visit. The photos will be reviewed by two independent reviewers, blinded to treatment assignment, who are clinical DFU experts, to assess wound healing. If there is discordance between the two reviewers, a third independent senior reviewer will provide the final assessment. To minimize missing data, participants who miss the final 20-week or the 2-week visit will be asked to send pictures of their feet. If >1 DFU is present, wound healing is based on the largest wound. If participants experience amputation or death before week 20, the ulcer will be classified as not healed.

#### Secondary outcomes

this RCT included a broad range of secondary outcomes, in line with the RE-AIM framework^28^. These secondary outcomes include metrics related to DFU healing, healthcare utilization, services accessed, patient reported outcomes, social determinants of health, and cost effectiveness.

##### DFU healing metrics

the US FDA defines complete wound healing as “skin re-epithelialization without drainage or dressing requirements confirmed at two consecutive study visits 2 weeks apart” ^58^. Although the CARE-D-Foot-Nav program is not a wound-healing product and is not subject to US FDA requirements, we will report a secondary outcome of complete DFU healing 2 weeks (11 to 21 days) after the week-20 visit by taking pictures of the wounds for independent review. Additional wound outcomes include: (a) difference in wound area between baseline and week 20, (b) major amputations (defined as any amputation above the ankle) between hospital discharge and week 20, (d) major amputation and/or all-cause death between hospital discharge and week 20, (e) major or minor amputation (i.e., any lower-extremity amputation) between hospital discharge and week 20, (f) major or minor amputation and/or all-cause death between hospital discharge and week 20, and (g) time to wound healing.

##### Patient reported outcomes

we will administer surveys at baseline and week 20 to measure changes in overall quality of life (EQ-5D-5L), DFU-related distress (diabetic foot ulcer-short form^59^), social support (interpersonal support evaluation list shortened version^60^), and medical mistrust (trust in physician scale^61^).

##### Individual drivers of health outcomes

we will administer surveys at baseline and week 20 to measure changes in food insecurity, housing stability, and financial resource strain. We will also assess tobacco cessation rates by participant self-report and changes in alcohol use via changes in the AUDIT-C score^62^.

##### Changes in continuous glucose monitoring (CGM) device rates

we will report the number of participants who initiate CGM use during the 20-week post-hospital discharge period to assess if the CARE-D-Foot-Nav program improves access to this diabetes management technology.

##### 12-month outcomes

Following completion of the 20-week program, all participants will resume usual care. To understand residual effects of CARE-D-Foot-Nav, at the end of 52 weeks, we will evaluate (1) major amputation, (2) all amputations, (3) all-cause death rates, and (4) healthcare utilization among CARE-D-Foot-Nav and control participants. We will ascertain these outcomes with a final close out phone call and medical record review.

##### Healthcare utilization

attainment of the four pillars of multidisciplinary DFU care within 14- and 30-days of hospital discharge is a key healthcare utilization outcome. It is defined as attending outpatient visits with any provider managing a) diabetes b) wound care c) antibiotics for DFU infection if applicable, and (d) obtaining non-invasive PAD testing if applicable. We will measure these outcomes singly and in combination. Additional outcomes include all-cause and DFU-related emergency department visits and/or re-admissions 30 days and 20 weeks post-hospital discharge. We will define DFU-related emergency department visits and re-admissions by the presence or absence of DFU billing codes.

##### Services accessed

We will assess utilization of available resources at the end of the study by review of the EMR (e.g., nutrition clinic, tobacco cessation program) and patient self-report (e.g., food banks visited).

##### CARE-D-Foot-Nav program costs

To measure the program cost, navigators will maintain a participant-level log of time providing services, transportation costs, and costs to provide phones to participants who do not have one. In addition, we will track overall program costs to purchase wound care supplies and other items to improve DFU care such as pill boxes and point-of-care glucose measurement material, such as glucometers.

##### Peer support group attendance

We will log attendance to the CARE-D-Foot-Nav peer support groups.

##### Reach

to gain insight into the clinical trial results generalizability we will report a CONSORT diagram with reasons for exclusion and compare the characteristics of the clinical trial participants to those of hospitalized at Grady and Emory Healthcare hospitals who did not participate in the trial. Grady and Emory hospitals account for ≥60% of Atlanta’s hospital beds and ∼1,400 people are hospitalized per year in these hospitals.

#### Implementation outcomes

##### Program adherence

We will measure participant adherence to weekly encounters and assess how adherence impacts outcomes in the per protocol analysis. We will also assess rates of program dropout, defined as missing navigator encounters for ≥4 consecutive weeks.

##### CARE-D-Foot-Nav implementation consistency and integrity

The program core functions are described in Table 1. We will collect audio recordings for all encounters, and research staff will listen to the recordings to document which components were or were not discussed using binary outcome of yes/no, documenting the appropriate reason why a topic was not discussed. They will measure consistency across navigators and by participants characteristics, such as sex, race, and preferred language. If consistency is low (<90% of components on 2 or more encounters), we will retrain navigators and conduct follow-up assessments, as needed.

##### Qualitative methods to identify factors influencing implementation of program fidelity and acceptability

We will use the Consolidated Framework for Implementation Research (CFIR), a widely accepted framework to identify factors influencing implementation outcomes such as fidelity and acceptability^30^. CFIR includes 48 constructs that may act as factors influencing implementation, which are organized into five domains: (1) innovation characteristics (e.g., relative advantage); (2) outer setting (e.g., local attitudes and conditions); (3) inner setting (e.g., recipient- and deliverer-centeredness); (4) individual roles and characteristics (e.g., implementation facilitators) and (5) implementation process (e.g., accessing context). We will conduct semi-structured interviews with key program stakeholders: 60 program participants, 5 navigators, and 10 health care providers delivering DFU care. Interviews are well suited to examine individual views and experiences with the program, generating in-depth information about perceived constraints on access to, fidelity of, and acceptability of the program^121^. They also provide an opportunity for individuals to discuss information they are not comfortable sharing in a group, such as dislikes of the program or reasons for non-adherence. Trained personnel will conduct 1-hour long audio-recorded interviews using interview guides tailored to each stakeholder type. Domains and constructs from CFIR will guide the creation of interview questions and probes. All guides will be pilot tested and refined, as needed.

##### Details of participant interviews

To develop a highly relevant and appropriate participant interview guide, we will enlist input from participants who complete the CARE-D-Foot-Nav during its first year. Starting in the second year of the trial, we will recruit and purposively sample participants randomized to the CARE-D-Foot-Nav who participated in and did not participate in ≥75% of the weekly encounters (n=30 in each group; total n=60) after completion of the program period (week 20). We will also use purposive sampling to recruit information-rich participants with variable demographic and baseline characteristics and clinical outcomes (e.g., healers, non-healers). The sample size of 60 interviews should be sufficient to reach both code saturation (the point at which the range of issues has been identified) and meaning saturation (the point at which a rich understanding of issues has been developed)^63^. The interview guide will be designed to evaluate acceptability of the program, mainly participants’ likes and dislikes of the program. Interviews will be conducted in English or Spanish, depending on participants’ language preference.

##### Details of provider interviews

To understand provider perspectives on program acceptability, we will recruit 10 DFU providers (physicians, advanced practice providers,and wound care nurses) who are not part of the research team to participate in interviews after trial completion in year 5. We will purposively sample providers from different specialties involved in DFU care. Interviews will explore likes and dislikes of the program, such as how the program affected clinic flow and patient care.

##### Details of navigator interviews

We anticipate navigator retention of 2-3 years and, as a result, 4-5 navigators will rotate through the program during the 5-year study period. We will interview navigators as they exit the program to understand difficulties they face in maintaining fidelity to program components.

#### Economic outcomes

We will perform a rigorous economic evaluation of the CARE-D-Foot-Nav program, incorporating both a a cost-effectiveness analysis (CEA) and distributional cost-effectiveness analysis (DCEA). Analyses will be conducted from two perspectives: the health care sector perspective, capturing direct medical costs, and the societal perspective, capturing patient time costs, caregiving burden, transportation, productivity losses, and other nonmedical costs. Two time horizons will be evaluated. First, we will perform a short-term CEA alongside the trial, covering the 20-week intervention period. Second, we will develop long-term economic models to estimate costs and effects at 10-year, 20-year, and up to 40-year) horizons, enabling assessment of the program’s potential lifetime value.

### SAMPLE SIZE

We identified 5 studies published within the last 10 years that compared DFU healing between patients that did and did not receive multidisciplinary DFU care (table 3)^64–68^. All studies were observational and reported that multidisciplinary DFU care was associated with increased healing, with odds ratio ranging from 2.20 to 7.27. Based on a chart review of a convenience sample of patients with DFUs receiving care in our institution, we expect a 20-week healing rate of 27.5% among usual care participants, in line with US national-level healing data^69^. Assuming the same healing rate for this trial usual care group, adopting a two-sided chi-squared test with alpha=0.05, after accounting for 15% attrition rate, we would need 270 participants to achieve an 80% power to detect a positive effect of CARE-D-Foot-Nav on 20 week healing with the odds ratio of 2.20, the lowest odds ratio among recently published studies^64–68^. In a worst-case case, where the attrition is 25% in the control arm and 15% in the intervention arm, we will have 80% power to detect the effect size equal to the odds ratio of 2.32, which is in line with the healing odds ratio observed in the above-cited studies on multidisciplinary DFU care and DFU healing.

**Table 3:** Studies reporting on multidisciplinary care and DFU healing.

| Author (Year) | Participants who received multidisciplinary care (Yes vs No) | Healing rate with vs without multidisciplinary care (when healing was measured) | Healing odds ratio ↑ |
| --- | --- | --- | --- |
| Akturk (2021) | 271 vs 79 | 56.5% vs 35.4% (20-week) | 2.37 |
| Blanchette (2019) | 41 vs 11 | 73.2% vs 27.3% (1-year) | 7.27 |
| Huizing (2019) | 133 vs 104 | 45.9% vs 66.0% (unclear) | 2.29 |
| Peng (2022) | 71 vs 71 | 83.0% vs 62.5% (1-year) | 2.93 |
| Riaz (2019) | 7106 vs 888 | 89.1% vs 78.8% (unclear) | 2.20 |

Due to the limitations on how we estimated the 20-week healing rate among participants assigned to usual care arm (i.e., chart review of a convenience sample), we will consider updating our sample size based on the results of another trial our group is conducting in the same setting. Specifically, we are conducting a separate RCT to investigate the impact of continuous glucose monitors (CGM) on post-hospital discharge DFU healing (NCT06054659), where participants are randomized to real time CGM vs a blinded CGM (the control arm), and results are expected in mid 2026. We will consider revising the sample size estimates for this navigator trial if the healing rate among the CGM control participants are significantly different than 27.5%.

### STATISTICAL ANALYSIS

#### Primary outcome (20-week post-hospital discharge complete DFU healing)

We will compare healing rates between CARE-D-Foot-Nav and usual care groups using the Chi-squared test. The intention-to-treat analysis will include all randomized participants, and the modified intention-to-treat analysis will exclude participants who meet exclusion criteria after the randomization (e.g., amputation of ≥2 toes at the initial hospitalization). We will also perform a 100% and a 75% per-protocol analysis that excludes participants who do not complete 100% or ≥75% of navigator encounters, respectively. These per-protocol analyses include all usual care participants.

We will further conduct logistic regression to estimate the group difference in the healing rate while adjusting for potential confounders, such the Society for Vascular Surgery Wound, Ischemia, foot Infection (WIfI) stage^34^. We will apply standard model selection (e.g., stepwise selection) and diagnostic procedures (e.g., Hosmer-Lemeshow test) to determine the form of the final model.

#### Secondary outcomes

We will use the Chi-squared test for binary secondary outcomes. For continuous secondary outcomes, we will assess the group difference in the outcome using the nonparametric Wilcoxon test or t-test and conduct linear regression to adjust for potential confounders. For binary secondary outcomes, we will follow the analysis plan outlined for the primary outcome. For secondary outcomes that are counts, we will compare them between the CARE-D-Foot-Nav and usual care groups using Mann-Whitney tests and perform Poisson regression or Negative Binomial regression to account for potential confounders. Some secondary outcomes are repeated measures outcomes, such as HbA1c measured at baseline and week 20. For such an outcome, we will first perform cross-sectional analyses that compare the outcome between the two study groups at each given time point following the plans described above for a continuous, binary, or count outcome. Then we will perform repeated measures analyses based on linear mixed models or generalized linear mixed models depending on whether the outcome measured at each time point is continuous or discrete. We will analyze the time to wound healing by using Kaplan Meier curves to estimate its distribution overall or in each study group. We will compare time to wound healing between the CARE-D-Foot-Nav and usual care groups using the nonparametric log-rank test. We will conduct Cox proportional hazard regression analyses to assess the group difference while accounting for potential confounders.

To analyze cost outcomes, such as health care utilization, we will follow the analysis plan outlined for the regular continuous outcome. To handle potentially skewed distribution of a cost outcome, we may consider utilizing transformed cost outcome or performing median or quantile regression. In all analyses, we will check the adequacy of fitted models with standard model checking procedures.

#### Missing data

To handle possible missing data, for example, due to patient dropout, we will assume that data are missing completely at random (MCR) and perform analyses based on the completely observed data. We will check the MCR assumption, for example, by evaluating the association of data missingness with patient characteristics via logistic regression. If a severe violation of MCR is identified, we will address missing data by applying multiple imputation or inverse probability weighting, based on appropriate modeling of data missingness.

#### Exploratory mechanism analysis using machine learning

To unravel the mechanisms underlying CARE-D-Foot-Nav effectiveness and guide its future refinement and broader implementation, we will employ advanced causal Artificial Intelligence (AI) techniques. This involves a nuanced analysis of the patient journey within the program, leveraging our novel hybrid causal AI method that integrates causal trees with causal forests ^70–73^. This analysis will focus on sequential data collected from baseline, during the intervention (e.g., use of CARE-D-Foot-Nav transportation resources), and at study completion (e.g., DFU healing). Our causal AI approach will elucidate two key aspects: the baseline characteristics that influence implementation traits, and the implementation factors that drive positive health outcomes. By identifying the profiles of individuals who benefit most and the critical success factors, we will deepen our understanding of the program’s operational dynamics and optimize target population identification, pinpointing program elements crucial for enhancing health outcomes.

#### Pre-specified sensitivity and subgroup analyses

We have two preplanned sensitivity analysis:

1-“Worst-case” analysis: all participants randomized to the CARE-D-Foot-Nav arm with missing outcomes are assigned a “not healed” outcome, and all participants assigned to the usual care arm with missing outcomes are assigned a “healed” outcome.

2-“Best case” analysis: all participants randomized to the CARE-D-Foot-Nav arm with missing outcomes are assigned a “healed” outcome, and all participants assigned to the usual care arm with missing outcomes are assigned a “not healed” outcome.

The subgroup analyses are based on (a) participant characteristics (b) wound severity metrics, (c) key comorbidities known to effect healing, (d) diabetes type and control, (e) individual drivers of health outcomes, (f) navigator assigned to the participant, and (g) program and hospital characteristics (Table 4).

**Table 4:**
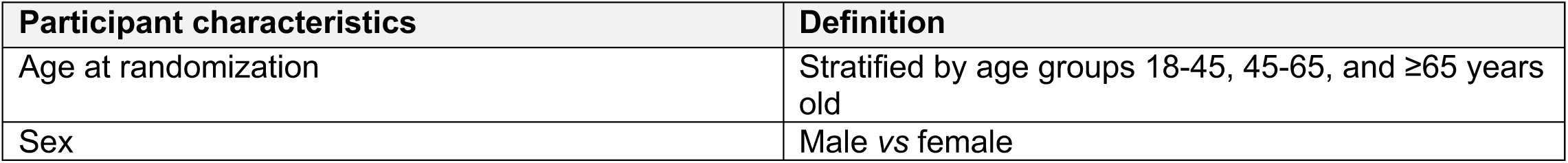
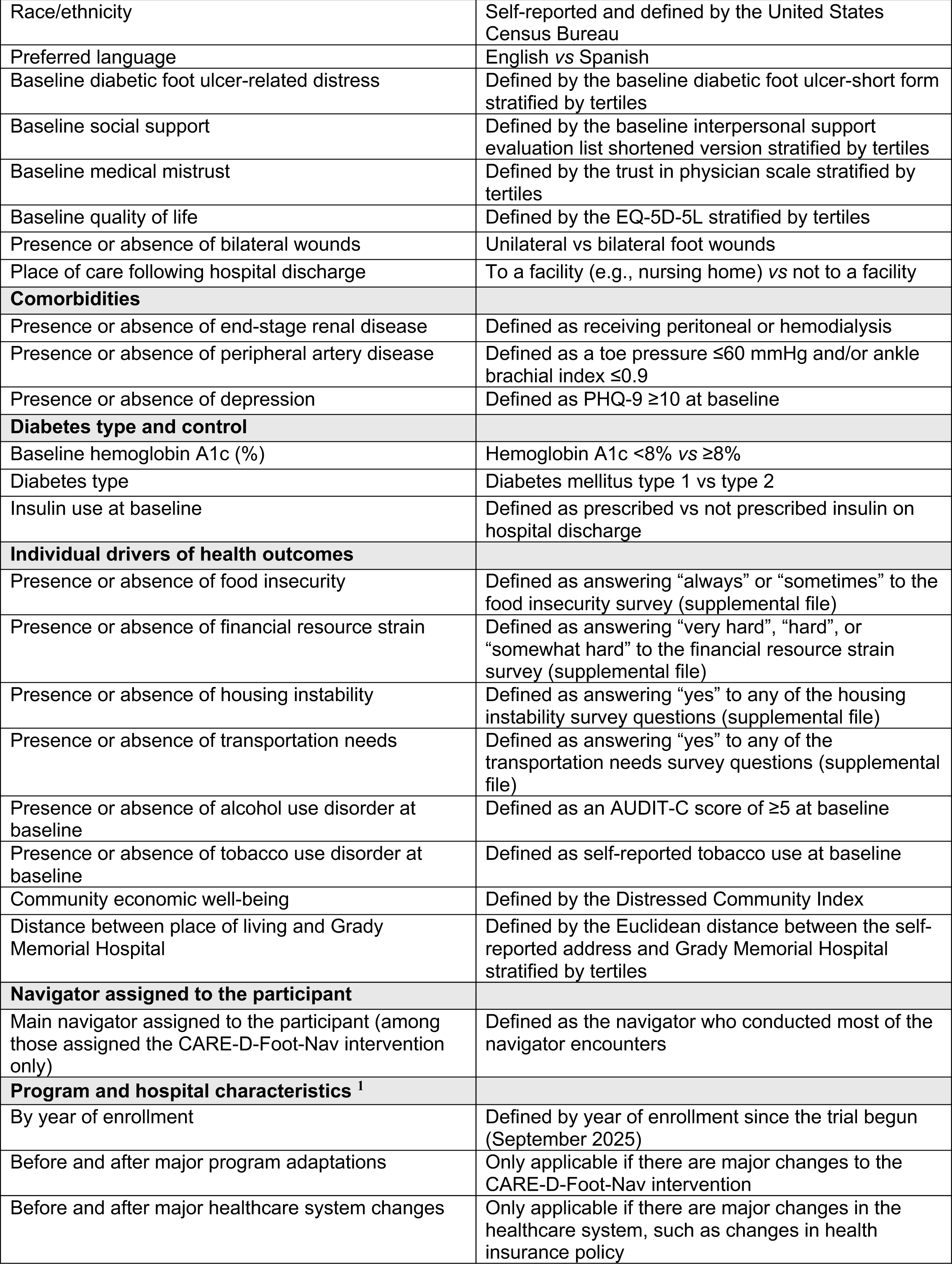
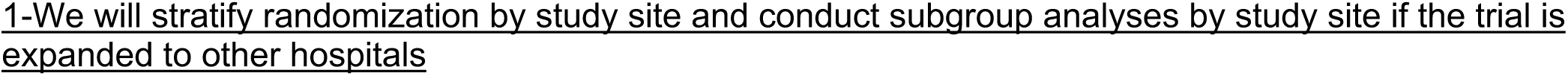
Pre-planned subgroup analyses.

| Participant characteristics | Definition |
| --- | --- |
| Age at randomization | Stratified by age groups 18-45, 45-65, and ≥65 years old |
| Sex | Male vs female |
| Race/ethnicity | Self-reported and defined by the United States Census Bureau |
| Preferred language | English vs Spanish |
| Baseline diabetic foot ulcer-related distress | Defined by the baseline diabetic foot ulcer-short form stratified by tertiles |
| Baseline social support | Defined by the baseline interpersonal support evaluation list shortened version stratified by tertiles |
| Baseline medical mistrust | Defined by the trust in physician scale stratified by tertiles |
| Baseline quality of life | Defined by the EQ-5D-5L stratified by tertiles |
| Presence or absence of bilateral wounds | Unilateral vs bilateral foot wounds |
| Place of care following hospital discharge | To a facility (e.g., nursing home) vs not to a facility |
| **Comorbidities** |  |
| Presence or absence of end-stage renal disease | Defined as receiving peritoneal or hemodialysis |
| Presence or absence of peripheral artery disease | Defined as a toe pressure $\leq 60$ mmHg and/or ankle brachial index $\leq 0.9$ |
| Presence or absence of depression | Defined as PHQ-9 $\geq 10$ at baseline |
| **Diabetes type and control** |  |
| Baseline hemoglobin A1c (%) | Hemoglobin A1c $< 8\%$ vs $\geq 8\%$ |
| Diabetes type | Diabetes mellitus type 1 vs type 2 |
| Insulin use at baseline | Defined as prescribed vs not prescribed insulin on hospital discharge |
| **Individual drivers of health outcomes** |  |
| Presence or absence of food insecurity | Defined as answering “always” or “sometimes” to the food insecurity survey (supplemental file) |
| Presence or absence of financial resource strain | Defined as answering “very hard”, “hard”, or “somewhat hard” to the financial resource strain survey (supplemental file) |
| Presence or absence of housing instability | Defined as answering “yes” to any of the housing instability survey questions (supplemental file) |
| Presence or absence of transportation needs | Defined as answering “yes” to any of the transportation needs survey questions (supplemental file) |
| Presence or absence of alcohol use disorder at baseline | Defined as an AUDIT-C score of $\geq 5$ at baseline |
| Presence or absence of tobacco use disorder at baseline | Defined as self-reported tobacco use at baseline |
| Community economic well-being | Defined by the Distressed Community Index |
| Distance between place of living and Grady Memorial Hospital | Defined by the Euclidean distance between the self-reported address and Grady Memorial Hospital stratified by tertiles |
| **Navigator assigned to the participant** |  |
| Main navigator assigned to the participant (among those assigned the CARE-D-Foot-Nav intervention only) | Defined as the navigator who conducted most of the navigator encounters |
| **Program and hospital characteristics <sup>1</sup>** |  |
| By year of enrollment | Defined by year of enrollment since the trial begun (September 2025) |
| Before and after major program adaptations | Only applicable if there are major changes to the CARE-D-Foot-Nav intervention |
| Before and after major healthcare system changes | Only applicable if there are major changes in the healthcare system, such as changes in health insurance policy |

1-We will stratify randomization by study site and conduct subgroup analyses by study site if the trial is expanded to other hospitals

### QUALITATIVE DATA ANALYSIS

All interview audio-recordings will be transcribed verbatim. Participant interviews in Spanish will be translated into English. All transcripts will be checked for accuracy and completeness and de-identified prior to analysis. Data will be analyzed using thematic analysis^74^. Textual data will be coded by two independent coders; inter-coder reliability will be assessed to identify any issues with the application of codes and promote consistent code application. Comparisons (e.g., adherence or non-adherence to ≥75% of CARE-D-Foot-Nav encounters) will be performed to understand differences or similarities in views and experiences across participants. All textual data will be managed and analyzed in MAXQDA 24.

### ECONOMIC ANALYSIS

We will provide decision-makers with crucial information regarding the program’s value, sustainability, and potential budgetary impact. We will conduct a cost-effectiveness analysis (CEA) and distributional cost-effectiveness analysis (DCEA) from both a health care sector and a societal perspective for two different time horizons: a short-term CEA alongside the trial (i.e., 20-week) and long-term (10-year, 20-year, and up to 40 years). We will follow recommendations of the Second Panel on Cost-Effectiveness in Health and Medicine ^75^. We will calculate the incremental cost-effectiveness ratio and net monetary benefit for implementing CARE-D-Foot-Nav compared to usual care. We will also extrapolate this analysis to settings outside the Grady hospital system to assess the cost-effectiveness and potential impact of implementing CARE-D-Foot-Nav on health outcome differences (i.e., DCEA) in these settings.

#### Cost-effectiveness analysis (CEA) alongside the trial

We will examine the 20-week cost-effectiveness of CARE-D-Foot-Nav.

##### Cost

When estimating costs from a health sector perspective, we will only include formal healthcare sector costs. Following a published method for service-line cost standardization^76^, we will extract line-level data for all EHR encounters and assign corresponding cost based on Medicare Fee Schedule: IPPS DRGs for inpatient facility services, OPPS/APCs for hospital outpatient/ED (with packaging/composite rules), MPFS for professional services (including telehealth/modifiers), CLFS for laboratory tests, ASP/DMEPOS for Part B drugs and CGM supplies, and NADAC (base case) or WAC (sensitivity) for Part D outpatient prescriptions (e.g., GLP-1RA, SGLT2 inhibitors, insulin pens). This benchmarking approach is transparent and reproducible, aligns with payer/health-system decision making, avoids site-specific charge idiosyncrasies, and enables cross-site comparability.

##### Quality-adjusted Life Year (QALY) Measurement

We will use the widely adopted EQ-5D to collect health utility data at baseline and 20 weeks. We will employ net-benefit regression to accurately assess the statistical uncertainty in the program’s cost-effectiveness estimation.

#### Long-term cost-effectiveness analysis (CEA)

For the long-term CEA of the CARE-D-Foot-Nav, we will use a microsimulation approach. Our team has previously developed and extensively validated the Building, Relating, Assessing, and Validating Outcomes (BRAVO) diabetes model, a person-level discrete-time microsimulation model ^77,78^. This model is widely recognized in the field of diabetes and chronic disease modeling and was used by many entities such as the CDC ^79,80^, the Institute of Clinical and Economic Review (ICER)^81^, and the ADA ^82^. The model will convert the short-term efficacy data of the CARE-D-Foot-Nav program obtained from the trial phase into lifetime health and cost outcomes. Health outcomes will encompass a variety of diabetes-related events beyond DFU and amputation, including macrovascular complications (such as myocardial infarction, stroke, and angina), microvascular complications (like retinopathy, blindness, and chronic kidney disease), hypoglycemia, and death. We will *adapt, validate, and calibrate* the BRAVO model to the trial’s population. We will use our previously developed cost equations ^83–85^ and QALY equations ^86,87^ to model the impact of diabetes complications on cost (health sector perspective) and health utilities, supplemented by additional estimates from relevant literature to get cost estimates from a societal perspective. We will extensively conduct sensitivity analyses to ensure the study’s robustness. We will also extend the simulation experiment settings to various populations beyond the Grady health system, utilizing external information sources such as US-based census data and state-level estimates. This approach will help inform the broader applicability of the program’s care model for future implementation across different settings.

#### Distributional cost-effectiveness analysis (DCEA)

DCEA is a state-of-art approach used in health economics and decision-making to incorporate considerations of distributional impacts alongside traditional CEA^88^. Our access to individual-level EHR data at Grady and the ability to perform individual-level microsimulation experiments over a lifetime offer a unique opportunity to study the DCEA of the CARE-D-Foot-Nav. In the proposed study, we will assess the distributional impact of CARE-D-Foot-Nav across various population subgroups and settings with various characteristics. We will also explore the efficiency trade-off across these populations. In addition, we will use machine-learning algorithms (e.g., random forest, gradient boosting) to identify potential subgroups that might have distinctive cost-effectiveness, and efficiency trade-off from the CARE-D-Foot-Nav program. Results from the DCEA will help decision-makers understand how CARE-D-Foot-Nav components affect different groups and assist in prioritizing interventions to achieve efficiency objectives.

### STUDY ORGANIZATION

This US NIH funded trial is led by the principal investigators (Fayfman and Schechter). The trial is approved by the Emory University Institutional Review Board and the Grady Memorial Hospital Research Oversight Committee. The steering committee is tasked with reviewing the trial progress and generate ideas to ensure success of all its operations. The data analysis team is led by Drs. Peng, Rhodes (qualitative analysis), and Shao (economic analysis). A data safety monitoring board will convene semiannually to review safety and data reports. Lastly, three external consultants who are DFU experts will serve as independent wound healing adjudicators and are blinded to the trial allocation arm.

## DISCUSSION

CARE-D-Foot-Nav is the first randomized controlled trial of a patient navigator for people with diabetic foot ulcers^24^. Our central hypothesis is that the CARE-D-Foot-Nav program will lead to higher healing rates compared to usual care by improving outpatient access to the 4 pillars of diabetic foot ulcer care ^8–10^. This trial will enroll 270 participants over 4 years; a relatively large sample size compared to available diabetic foot ulcer trials^69^. Aside from healing outcomes, the trial will evaluate patient-reported and economic outcomes to investigate the potential broader impact of the CARE-D-Foot-Nav program.

Using the RE-AIM implementation science framework, we will systematically evaluate implementation outcomes and generate generalizable evidence to inform the delivery of future patient navigator interventions. Robust fidelity reviews of patient encounters ensure alignment with the core functions of the intervention protocol, as well as support quality assurance efforts and prompt action throughout the trial if fidelity is found to be low (<90%). Additionally, we will conduct qualitative research guided by the CFIR to gain an in-depth understanding of implementation determinants (i.e., barriers and facilitators). Rich qualitative data on determinants will be essential for designing and testing implementation strategies to overcome identified barriers and promote identified facilitators in future research, ultimately informing the delivery and dissemination of this intervention in routine care settings.

The trial strengths include its high standards for rigor and reproducibility, including the RE-AIM and CFIR frameworks^28,30^, adherence to CONSORT guidelines^32^, and diabetic foot ulcer trial best practices^89^, such as having healing outcomes adjudicated by independent reviewers blinded to trial allocation arm. We will audio-record all navigator encounters to allow for a robust fidelity assessment. The trial’s main limitations are the single-center design and inclusion of participants who speak English and/or Spanish only, which may limit generalizability. However, we will use electronic medical databases to compare the characteristics of participants enrolled in the trial with those of persons hospitalized with diabetic foot ulcers in Atlanta, GA, who were not enrolled in the trial.

In conclusion, if the CARE-D-Foot-Nav patient navigator program proves to improve diabetic foot ulcer outcomes, it could change how we care for people with this disease and decrease preventable limb loss.

## Data Availability

All data produced in the present study are available upon reasonable request to the authors

